# The Causal Association between Gut Microbiota and Pancreatic Cancer: A Two-sample Mendelian Randomization Analysis in European and East Asian Populations

**DOI:** 10.1101/2024.04.23.24306212

**Authors:** Yinbo Xiao, Xiang Li, Long Zou, Daniel M. Wall, Zhiyong Liang

## Abstract

**Background and Aim:** Alterations in the gut microbiota strongly correlate with the onset of pancreatic cancer (PC). However, any causal relationship between gut microbiota alterations and PC risk remains unknown.

**Methods:** We comprehensively investigated PC-related microorganisms in European and East Asian populations through the application of Mendelian randomization (MR). The PC genome-wide association study (GWAS) databases for European and East Asian individuals were acquired from the UK and Japanese Biobanks, respectively. Primary analytical methods, including the inverse variance weighted (IVW) method, weighted median, Maximum likelihood method and MR PRESSO, were employed to estimate the potential causal association between gut microbiota and PC. Additionally, we performed sensitivity analysis and reverse MR analysis.

**Results:** By IVW method, overall 17 bacterial taxa were identified with potential causal correlations to PC. The PC-associated gut microbiota signatures varied across different populations. Among these, 4 specific taxa exhibited potential causality with PC, with statistical significance in all four MR methods. Specifically, the *Alcaligenaceae* family was identified as protective, while genus *Sutterella*, order *Bacilliales* and genus *Enterohabdus* were associated with increased risk of PC. Among the European population within the UK biobank, the *Alcaligenaceae* family, genus *Sutterella*, and order *Bacillales* were connected to PC, while genus *Enterohabdus* was linked to PC in the Japanese cohort.

**Conclusion:** Our study implicates certain members of the gut microbiota in PC onset based on genetics. Further investigations of the gut-pancreas axis may lead to the development of novel microbiome targeted prevention strategies for PC.

## Introduction

Pancreatic cancer (PC), especially pancreatic ductal adenocarcinoma, is one of the most lethal malignancies worldwide [1]. With an estimated 49,830 deaths attributed to PC in 2022, it remains the fourth most common cause of cancer-related deaths in the US [2]. Moreover, it is predicted to become the second leading cause of cancer-related deaths by 2030 in the US [3]. The incidence and mortality rates of PC are continuously increasing year by year, unfortunately with minimal progress in the overall survival rate [4]. Thus, it is urgent to gain a deeper understanding of the biological mechanisms underlying PC carcinogenesis, potentially leading to novel treatment and management strategies for PC to reduce such public health burden.

The role of the gut microbiome in PC development, known as the microbiome-pancreas axis, has gained significant attention [5]. Microbiota from the gut can enter the pancreas via the circulatory system or pancreatic duct [6, 7]. Numerous research studies have revealed that gut microbiota alterations are involved in the advent of PC [8–11], even though the underlying molecular mechanisms remain unclear. Moreover, some epidemiological studies have demonstrated that the risk of PC is positively correlated to the abundance of some gut microbiota, such as *Aggregatibacter actinomycetemcomitans*, *Porphyromonas gingivalis* and *Helicobacter Pylori* [10, 12, 13]. These studies indicated that the gut microbiota could serve as biomarkers for PC in clinical practice and be utilized for early detection of PC and prognosis of outcomes. However, a causal role for the gut microbiota in PC has not been established, due to confounding factors and the potential for reverse causality [14–16]. Mendelian randomization (MR) is an epidemiological method whereby genetic variants are employed as instrumental variables (IVs) to determine the causal relationship between risk factors and disease outcomes [17, 18]. The use of genetic variants is advantageous as they are randomly distributed during conception, reducing the impacts of confounding factors and eliminating reverse causation bias. Consequently, MR analysis is less susceptible to the influence of environmental and self-adopted confounding factors [19]. In this study, MR offers a valuable approach to estimate the causal link between the gut microbiota and the risk of PC.

Using MR analysis, previous studies have identified the causal relationships between the gut microbiome and several cancers, including liver cancer [20], colorectal cancer [21], and lung cancer [22]. However, any causal association between the gut microbiota and PC is still unclear. Furthermore, most MR studies are largely derived from European populations, while there is a limited number of studies employing MR analysis in non-European cohorts. Here, we performed a two-sample MR analysis to evaluate the association between the gut microbiome and PC risk among both European and East Asian populations. Our study can enhance the theoretical basis for the gut-pancreas axis leading to novel insights into the predictors of PC as well as potential treatment targets.

## Methods

### Exposure data

Genetic variants related to the gut microbiota composition were acquired from the large-scale genome-wide association study (GWAS) [23]. The gut microbiota was profiled by targeting three variable regions V1–V2, V3–V4 and V4 of the 16S rRNA gene. The meta-analysis encompassed a total of 18,340 participants derived from 24 cohorts originating from several countries including the United States, Canada, Israel, South Korea, Germany, the United Kingdom. Following adjustment for age, gender, technical covariates, and genetic principal components, the quantitative microbiome trait loci analysis yielded a total of 211 GWAS summary statistics associated with microbial taxa. These were 9 phyla, 16 classes, 20 orders, 35 families (including 3 families with unknown classification), and 131 genera (with 12 genera of unknown classification). The summary data is available for download from https://mibiogen.gcc.rug.nl/.

### Outcome data

The UK Biobank and the Japan Biobank were used to investigate the causal relationship between the microbiome and pancreatic cancer. The cohort data from the UK Biobank of PC involved 589 PC cases and 393372 healthy control. Summary analysis statistics are available from the Lee Lab (https://www.leelabsg.org/resources). The summary GWAS data of the Japan Biobank included 442 PC cases and 195745 healthy controls. Data was acquired from the IEU Open GWAS project (https://gwas.mrcieu.ac.uk/datasets/).

### Assumptions

The schematic representation of the investigation is depicted in **Figure 1**. In this study, the gut microbiota was considered as the exposure, while PC was regarded as the outcome. Three fundamental assumptions were necessary for a proper MR study. First, the genetic variants designated as IVs were strongly related to the exposure. Second, the relationships between genetic variations and outcomes were not influenced by any other confounding variables. Third, it should be noted that the impact of genetic variations on the outcome was only mediated by their influence on the specific exposure under investigation. It meant that no occurrence of horizontal pleiotropy was shown between the genetic variants and the outcome.

**Figure 1.**
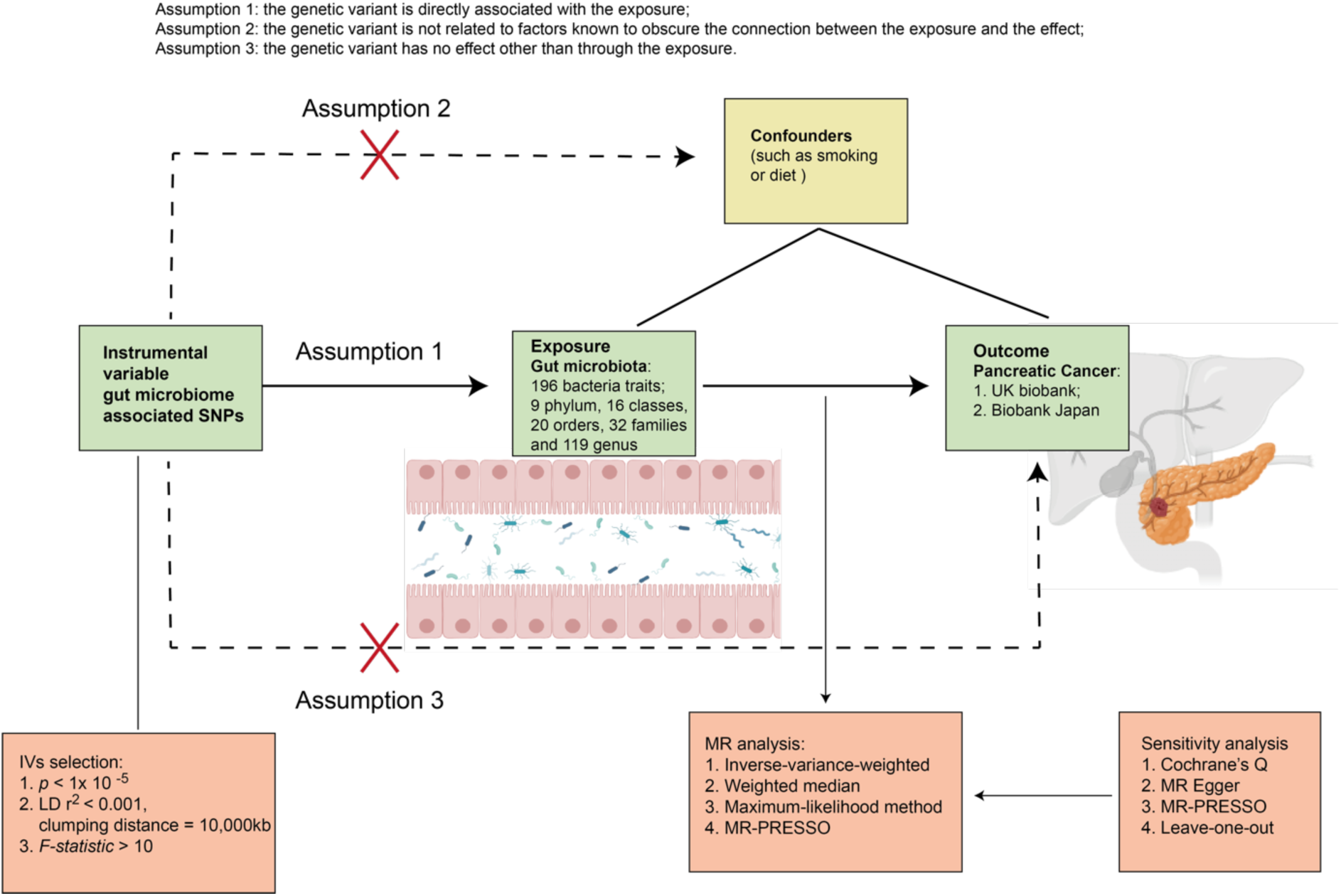
The study design and the overall workflow.

### Instrumental variable selection

Excluding 15 unknown classifications, a total of 196 taxa at five different levels (phylum, class, order, family, and genera) were conducted as the exposure datasets. Potential IVs were selected by single nucleotide polymorphisms (SNPs) with a less stringent significant association at a threshold of *P* < 1.0 × 10^−5^. The approach was employed to augment the pool of SNPs accessible for conducting sensitivity tests, as demonstrated in prior studies [23, 24]. Independent SNPs were clumped as IVs based on linkage disequilibrium (LD) R^2^ < 0.01 and clumping distance equal to 10,000 kb. The strengths of the IVs were estimated by F statistics. Specifically, the extent to which the IVs accounted for variance was computed for each exposure. F statistics were calculated with the following equation, r^2^ * (N – 1 – k)/ [(1 – r^2^)* k], where r^2^ was the variance explained, N was the sample size and k was the number of IVs [25]. To calculate F statistics for independent IVs, k was equal to 1. Finally, independent IVs with F-statistics below 10 were deemed to be weak IVs and therefore were eliminated from the analysis. To identify whether selective IVs were associated with confounders, PhenoScanner was applied to exclude IVs significantly associated (*P* < 5x10^-8^) with potential confounders (i.e., obesity, smoking, diet, or other diseases).

### Mendelian randomization analysis

MR analysis was conducted in R using TwoSampleMR package version 0.5.7 [26]. Selected IVs from different gut microbiota taxa were combined with the PC outcome SNPs dataset. To ensure the effects of SNPs on the exposure corresponding to the same allele as the effects on the outcome, shared SNPs were harmonised across exposure and outcome databases. At least 3 shared SNPs available between exposure and outcome were then selected for further MR analysis. To evaluate causal estimates between the gut microbiota and the risk of PC, MR causality tests were performed using four different approaches: inverse-variance-weighted (IVW) method, weighted median, maximum-likelihood method and MR-PRESSO [27]. In particular, the IV method of estimation was fundamentally a meta-analysis technique which was operated under the assumption that IVs had a causal impact on the outcome solely through the exposure, rather than through any other pathways [28]. The weighted median estimator provided valid estimations of causal effect when no more than 50% of the information was from invalid instruments. MR PRESSO was employed to estimate the pleiotropy, which corrected the estimation by eliminating outliers from the IVW model. Therefore, the presence of a causal relationship was determined when a statistically significant *P* value (*P* < 0.05) was obtained from any of the four methods used in the MR analysis.

### Sensitivity analysis

After MR analysis, sensitivity analysis was performed to evaluate potential heterogeneity and pleiotropy. The Cochran’s Q statistics were employed for heterogeneity analysis. When the p-value of Q statistics was less than 0.05, it might be interpreted as evidence of heterogeneity. MR-Egger intercept, as well as MR-PRESSO test, were conducted to monitor the potential horizontal pleiotropy. An insignificant *P* value (*P* > 0.05) in the MR-Egger intercept test was defined as the absence of pleiotropy. The MR-PRESSO test was also conducted to examine pleiotropic biases and address the pleiotropic effects by eliminating outliers. MR analysis report was not supported by the MR PRESSO outliers-adjusted test (*P* > 0.05) representing substantial pleiotropy. In addition, leave-one-out analysis was performed to determine if the causal estimates were biased by any one single SNP. Leave-one-out analysis was able to identify one SNP driving the signal when all, but one leave-one-out configuration had *P* < 0.05.

### Reverse MR analysis

To determine whether PC had any causal impact on the identified significant microbiota, a reverse MR analysis was conducted (PC as the exposure and the identified significant gut microbiota as the outcome), by using SNPs that were strongly associated with PC as IVs (p < 5x10^-6^). Causal tests based on the MR framework were then conducted, following the same methodology as described in the section “Mendelian randomization analysis”. To eliminate intricate causality possibility, we also excluded any results in which the *P*-value of reverse MR was less than 0.05.

## Results

### Selection of instrumental variables

IVs were sorted by p < 1x10^-6^. After excluding unknown bacterial genera or ones containing less than three IVs, a total of 119 bacterial genera were used as exposure datasets. The F-statistics of IVs were more than 10, suggesting that there was no evidence of weak instrument bias. Details about the selected IVs for 119 genera were shown in **Supplementary Table S1**.

### Association of specific members of the gut microbiota with PC

In the MR analysis, 17 bacterial genera were genetically predicted associated with the risk of PC in the IVW method of MR analysis. Specifically, there were 11 genera (class *Bacyeroidia*, family *Alcaligenaceae*, family *Veillonellacease*, genus *Bilophila*, genus *Eggerthella*, genus *LachnospiraceaeUCG004*, genus *LachnospiraceaeUCG010*, genus *Parasutterella*, genus *Sutterella*, order *Bacillales*, and order *Bacteroidales*) in European ancestry from the UK Biobank (**Table 1**), while 6 genera (class *Atinobacteria*, family *Christensenellaceae*, genus *Ruminococcusgnavus* group, genus *Enterohabdus*, genus *Ruminococcus1*, order *Burkholderiales*) were identified in East Asian ancestry from the Japan Biobank (**Table 2**). The scatter plots of IV potential effects on PC versus gut microbiota in the European and East Asian populations were demonstrated in **Supplementary Figure S1 and S2,** respectively. In order to identify the strongest evidence of significant risk factors between any microbial taxa and PC, 17 significant bacterial genera were valued in 4 different MR analysis methods (IVW, weighted median, Maximum likelihood, and MR PRESSO). The most significant risk factors were required to achieve a *p*-value below 0.05 in all four distinct techniques of MR analysis. Hence, four microbial taxa were identified that fulfilled these requirements, including three taxa (family *Alcaligenaceae*, genus *Sutterella*, and order *Bacillales*) being associated with PC in the UK Biobank cohort, and one taxon (genus *Enterohabdus*) being associated with PC in Japan Biobank cohort (as shown in **Table 1** and **2**). Meanwhile, the IVW, weighted median, Maximum likelihood, and MR PRESSO, all 4 analysis methods produced similar casual estimates for magnitude and direction. In detail, the family *Alcaligenaceae* (OR = 0.5, 95% CI = 0.29-0.86, *p* = 0.011, IVW) had a protective effect on PC in European populations, while genus *Sutterella* (OR = 2.25, 95% CI = 1.27-3.99, *p* = 0.005, IVW) and order *Bacillales* (OR = 1.60, 95% CI = 1.18-2.16, *p* = 0.002, IVW) were associated with a higher risk of PC. The findings from the Japanese cohort indicated a heightened likelihood of developing PC correlated with the presence of the genus *Enterohabdus* (OR = 2.38, 95% CI = 1.40-4.04, *p* = 0.001, IVW).

**Table 1.**
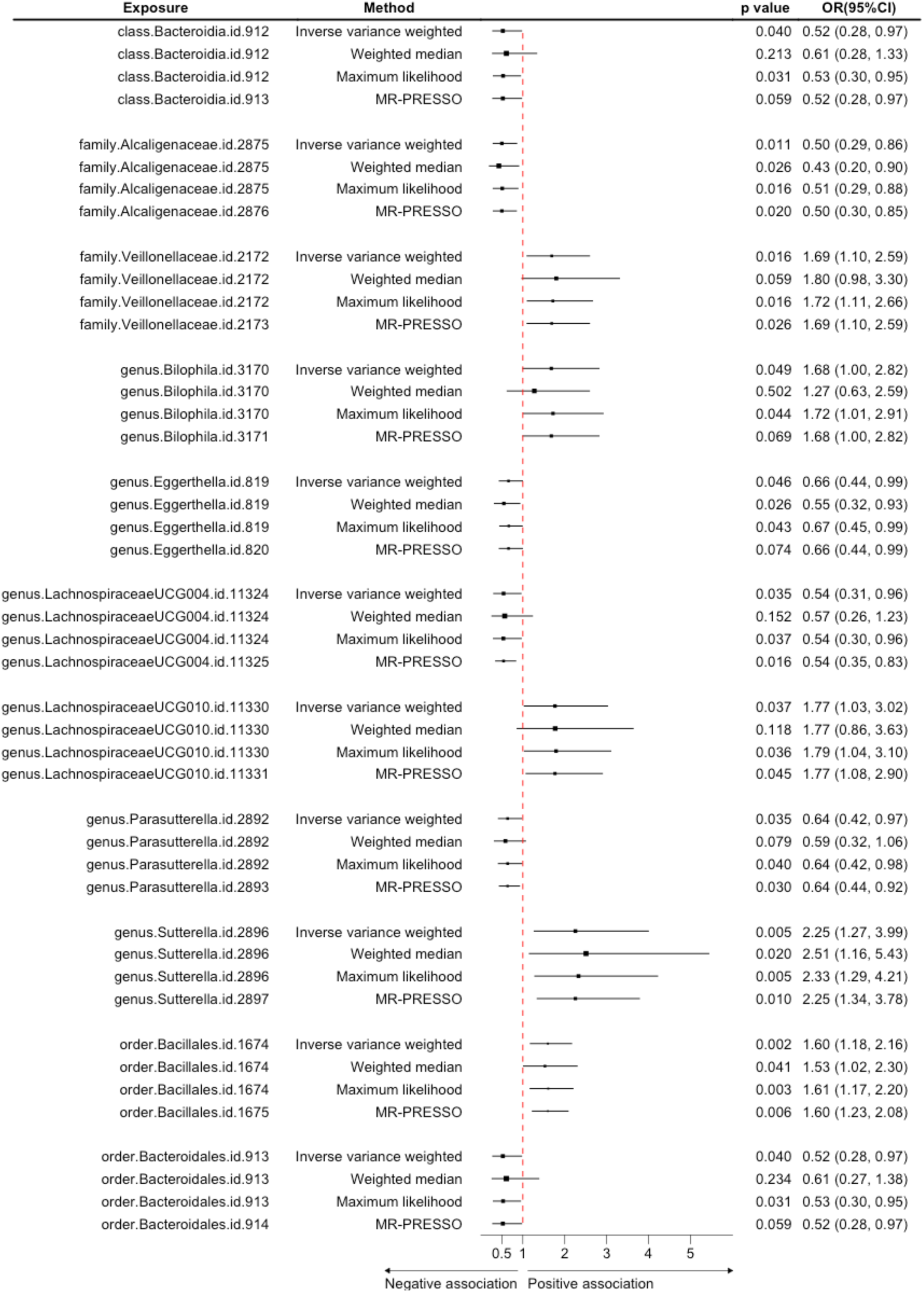
Causal associations of the gut microbiota with pancreatic cancer risk in the European population.

**Table 2.**
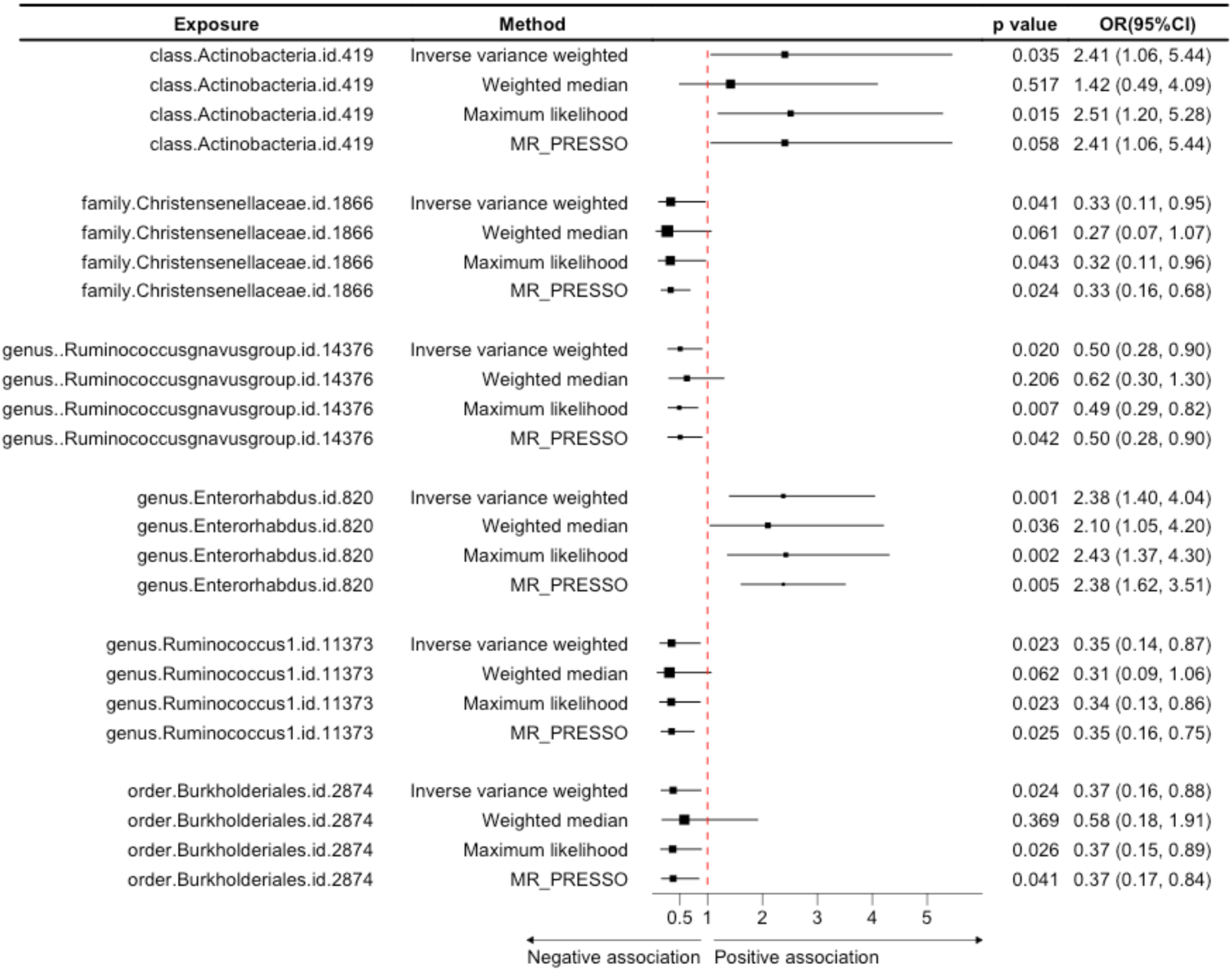
Causal associations of the gut microbiota with pancreatic cancer risk in the East Asian population.

### Sensitivity Analyses

The Cochran’s Q statistics for all 17 significant risk factors of gut microbiota in PC showed no significant heterogeneity (*p* > 0.05) (**Supplementary Table S2**). Meanwhile, no evidence of horizontal pleiotropy for gut microbiota in PC with *p* > 0.05 was demonstrated by the MR-Egger regression intercept approach (**Supplementary Table S3**). The heterogeneity of 17 significant risk factors of PC was also revealed by the MR-PRESSO global test, and the analysis revealed no outliers in the results (**Supplementary Table S4**). Moreover, leave-one-out analysis exhibited no significant difference in causal estimations of all 17 bacterial genera on PC (**Supplementary Figure S3 and S4**). No association between Pancreatic cancer and the following taxonomic groups was observed in the reverse MR analysis of the UK Biobank data: class *Bacyeroidia*, family *Alcaligenaceae*, family *Veillonellacease*, genus *Bilophila*, genus *Eggerthella*, genus *LachnospiraceaeUCG004*, genus *LachnospiraceaeUCG010*, genus *Parasutterella*, guens *Sutterella*, order *Bacillales*, and order *Bacteroidales* (**Table 3**). As there were few IVs identified from the Japanese Biobank, only 7 IVs were selected even when the cut-off *p*-value was set as 1x10^-5^. Therefore, we could not perform reserve MR analysis between PC and the gut microbiome in the population from Eastern Asia. Detailed information on the IVs (*P* value < 5x10^-6^) used in the reverse MR analyses was shown in **Supplementary Table S5**.

## Discussion

Our study used MR analysis to offer valuable insights into the potential causal relationship between gut microbiota and PC. IVW estimates suggested that within the European population, class *Bacteroidia*, family *Alcaligenaceae*, genus *Eggerthella*, genus *LachnospiraceaeUCG004*, genus *Parasutterella*, order *Bacteroidales* were related to the reduced risk of PC, while family *Veillonellaceae*, genus *Bilophila*, genus *Lanchnospiraceae UCG010*, genus *Sutterella*, and order *Bacillales* were positively related to the risk of PC. In the East Asian population, several gut microbiota members were identified to be related to the reduced risk of PC, including family *Christensenellaceae*, genus *Ruminococcusgnavus group*, genus *Ruminococcus 1*, and order *Burkholderiales*, while the level of class *Actinobacteria* and genus *Enterorhabdus* were positively related to the PC. These findings not only contribute to the advancement of our knowledge of microbiota in the development of cancer, but also highlight the importance of ethnicity in the risk of PC.

Based on the PC GWAS database from the UK biobank and the Japan biobank, our study indicated that the PC-associated gut microbiome displayed widespread regional differences. Furthermore, a recent meta-analysis conducted on the Finnish biobank data identified a distinct group of bacteria associated with PC. However, these findings varied from the results obtained from biobanks in the UK and Japan. [29, 30]. All this evidence suggested that the PC-related gut microbiota signatures varied across different populations. Previous studies demonstrated strong associations between race (European, African, Asian) and different genera abundances in most cancer types [31]. The incidence of PC is greater in Europe than in East Asia [32]. The disparity may be associated with gut microbiome differences. However, the microbiome profiles among various racial groups in PC, are not well understood. The different abundant bacteria might be attributed to various dietary patterns among individuals from distinct regions. It is noteworthy that the diversity in human gut microbiome composition between ethnic groups manifests as early as three months after birth [33]. Hence, diverse geographic populations should be considered in the microbiota-based disease models [34]. The identification of unique gut microbiota linked to PC in the UK and Japan provide an opportunity to further conduct country-specific studies on the development of PC.

The findings generated from our MR analysis were consistent with the previously published results obtained from 16s rRNA sequencing of microbiota in PC patients. The clinical PC faecal microbial profile by 16s rRNA sequencing reported that *Eggerthella* and *Parasutterella* were significantly decreased in PC patients compared with healthy control [35]. Another faecal microbiome signature in PC patients was characterised by a decreased presence of bacterial families commonly in the healthy gut, namely *Ruminococcaceae* and *Lachnospiraceae*; and an increased presence of *Veillonellaceae* [36]. Additionally, a comparison of the relative abundances of each microbial species revealed that *Sutterella Wadsworthensis* and *Bilophila* were significantly enriched, while *Bacteroildes Rodentium* was significantly decreased in PC as compared with healthy controls [37, 38]. Thus, the two-sample Mendelian randomization study is thought to provide a convincing approach for evaluating the relationships between gut microbiota and PC, as it is compatible with faecal 16s sequencing results.

According to our findings, a decrease in the abundance of *Lachnospiraceae* and *Ruminococcaceae*, as well as an increase in *Veillonellaceae*, were associated with the PC risk. Strikingly, a similar profile of altered gut microbiota was also exhibited in the patients diagnosed with cirrhosis [39, 40]. In a linkage study in southern England, elevated risks of PC were identified to be related to earlier liver diseases, such as alcoholic cirrhosis, primary biliary cirrhosis and unspecified cirrhosis [41]. Combined with our results, new research will be proposed that a potential role of altered gut flora in cirrhosis patients could contribute to the increased susceptibility to PC. Besides, it is well acknowledged that obesity substantially elevates the risk of PC; however, the underlying mechanisms connecting the two remain poorly understood [42]. The genus *Eggerthella,* as a protective factor for PC, exhibited a significantly lower abundance in obese individuals compared to non-obese ones [43].

Conversely, the genus *Sutterella* as a risk factor for PC, exhibited an increase in obese people [44, 45]. Given the similarity of the microbiota profiles between the obese population and PC patients, it is reasonable to assume that the microbiota alteration, such as the metabolic changes linked to the *Eggerthella* and *Sutterella* [46], could mediate the mechanism by which the obesity initiates the development of PC. Taken together, our study can provide novel insights into the relationships between PC, the microbiome and other risk factors, which could enhance our knowledge of PC development.

The therapeutic techniques aiming at the cancer-associated microbiome have been conducted in clinical trials. In this context, our study found that *Ruminococcus* and *Lachnospiraceae* were severed as the protective role of PC, suggesting novel and potential treatment targets for gut microbiome-based therapy. Hester et al. [47] demonstrated that the consumption of a substantial quantity of dietary fibre led to the production of a significant level of butyrate production by several bacterial family, such as *Lachnospiraceae* and *Ruminococcaceae*, which had preventive properties against the development of colon cancer. The utilization of probiotics in healthy individuals has been found to inhibit the development of colon carcinoma by increasing the number of *Ruminococcus* species and *Clostridiales* bacteria [48, 49]. Therefore, the use of probiotics containing *Ruminococcus* could have promise as a novel therapeutic approach for mitigating the onset of PC. On the other hand, based on our findings, other PC-associated microbiota are worthy of further investigation, making this a promising direction for targeted gut microbiome-based therapy for PC.

Our study has several limitations. First, our research included a total of 119 microbial taxa; however, we did not investigate potential causal associations at the species level. Second, MR analysis is a computer-based correlation analysis between gut microbiota and PC, without explaining the underlying process. It would be advantageous to validate the outcomes via functional experiments. Third, the present study included people of European and East Asian descent, perhaps restricting the generalizability of the findings to other populations.

In conclusion, this study identified several candidate bacteria that have potential association with PC. Variations in the PC-associated gut microbiota signatures are evidenced with geographical location, which may explain the disparity of PC incidence across nations. The identification of PC-associated gut microbiota provides the foundation for the exploration of novel microbiota-targeted therapy for PC. Further studies are needed to better characterise the potential role of these gut microbiota in the pathogenic mechanisms of PC.

## Data Availability

The gut microbiota summary data is available for download from https://mibiogen.gcc.rug.nl/.
European biobank of Pancreatic cancer Summary analysis statistics are available from the Lee Lab (https://www.leelabsg.org/resources).
Pancreatic cancer in Japan Data was acquired from the IEU Open GWAS project (https://gwas.mrcieu.ac.uk/datasets/).

https://www.leelabsg.org/resources

https://gwas.mrcieu.ac.uk/datasets/

---

**Supplementary Figure 1.**
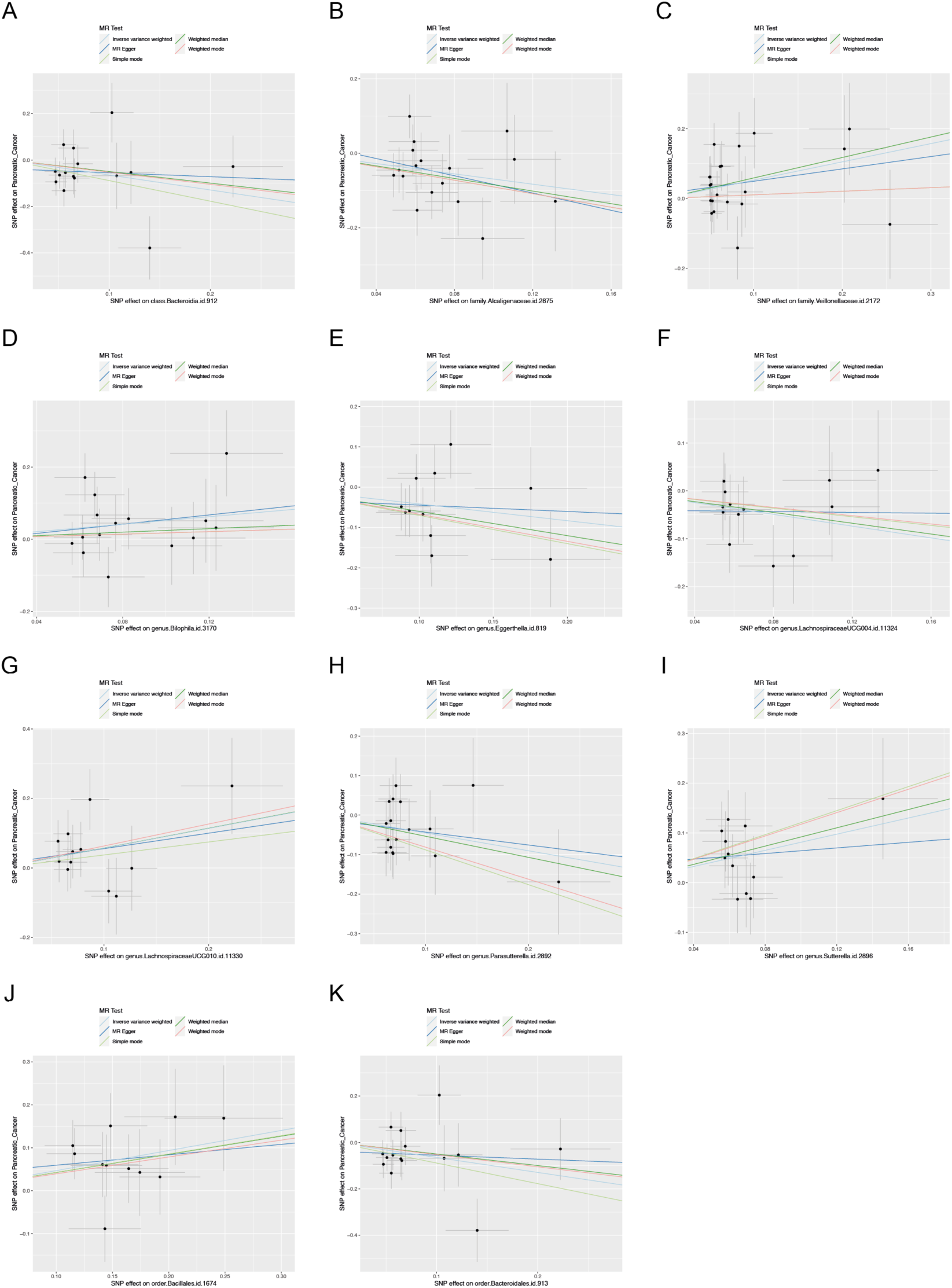
Scatter plot of the association between gut microbiota and pancreatic cancer based on UK biobank database. (A) class *Bacteroidia*; (B) family *Alcaligenaceae*; (C) family *Veillonellaceae*; (D) genus *Bilophila*; (E) genus *Eggerthella*; (F) genus *LachnospiraceaeUCG004*; (G) genus *LachnospiraceaeUCG010*; (H) genus *Parasutterella*; (I) genus *Sutterella*; (J) order *Bacillales*; (K) order *Bacteroidales*

**Supplementary Figure 2.**
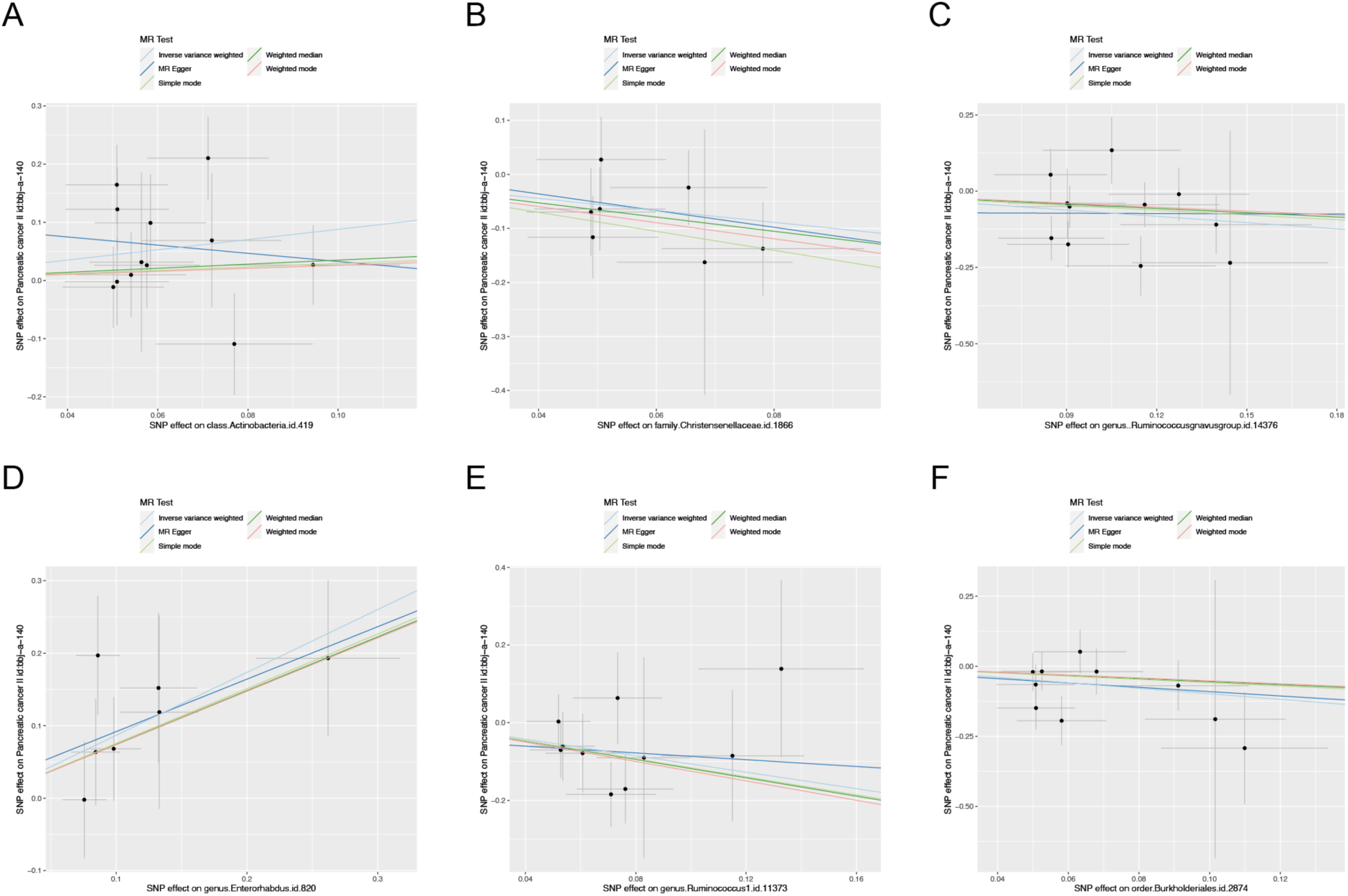
Scatter plot of the association between gut microbiota and pancreatic cancer based on Japan biobank database. (A) class *Actinobacteria*; (B) family *Christensenellaceae*; (C) genus *Rumminococcusgnavus*group; (D) genus *Enterorhabdus*; (E) genus *Ruminococcus1*; (F) ordr *Burkhoderiales*

**Supplementary Figure 3.**
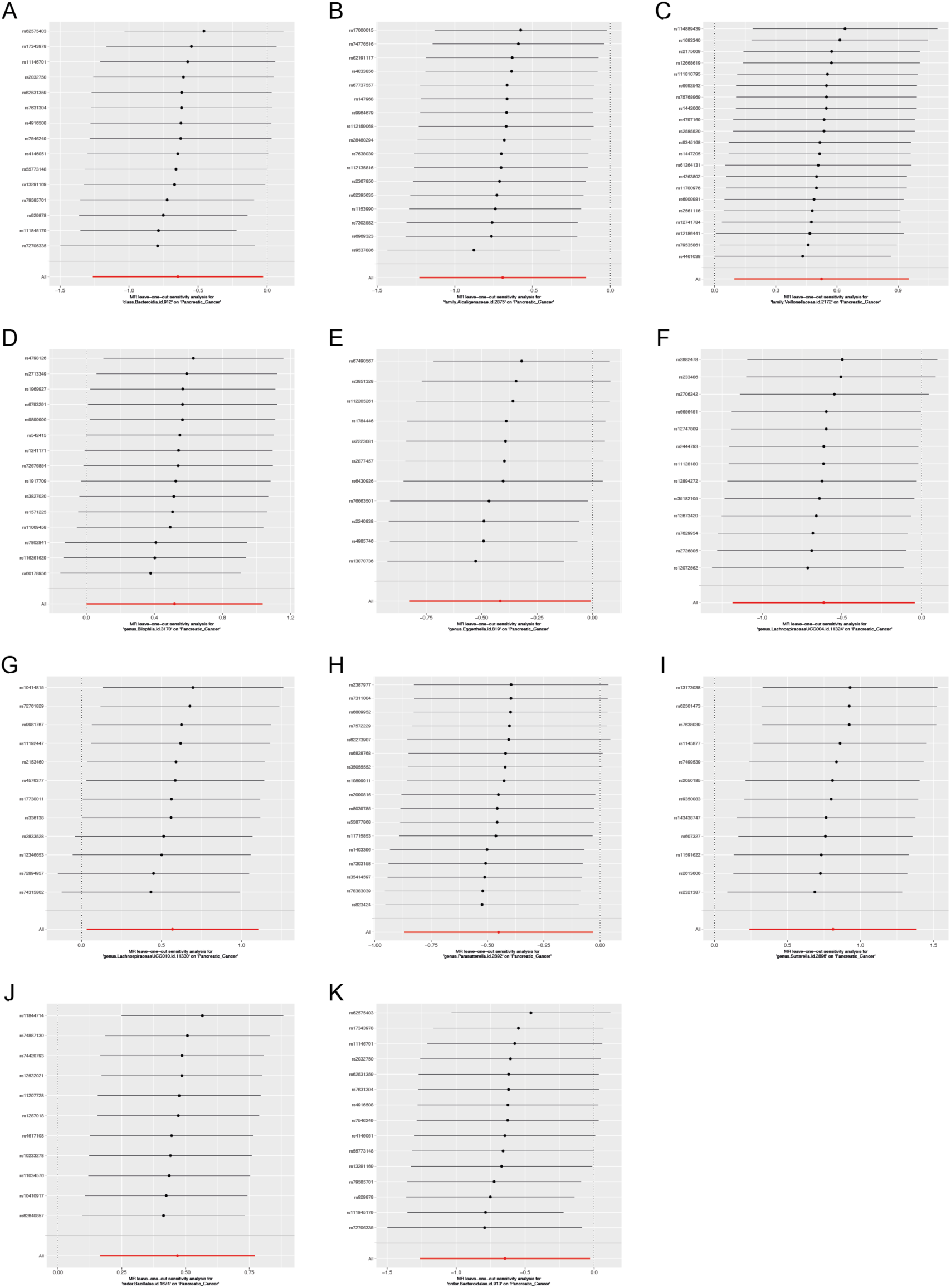
Leave-one-out sensitivity analysis for the association between genetically predicted gut microbiota and pancreatic cancer based on UK biobank database. (A) class *Bacteroidia*; (B) family *Alcaligenaceae*; (C) family *Veillonellaceae*; (D) genus *Bilophila*; (E) genus *Eggerthella*; (F) genus *LachnospiraceaeUCG004*; (G) genus *LachnospiraceaeUCG010*; (H) genus *Parasutterella*; (I) genus *Sutterella*; (J) order *Bacillales*; (K) order *Bacteroidales*

**Supplementary Figure 4.**
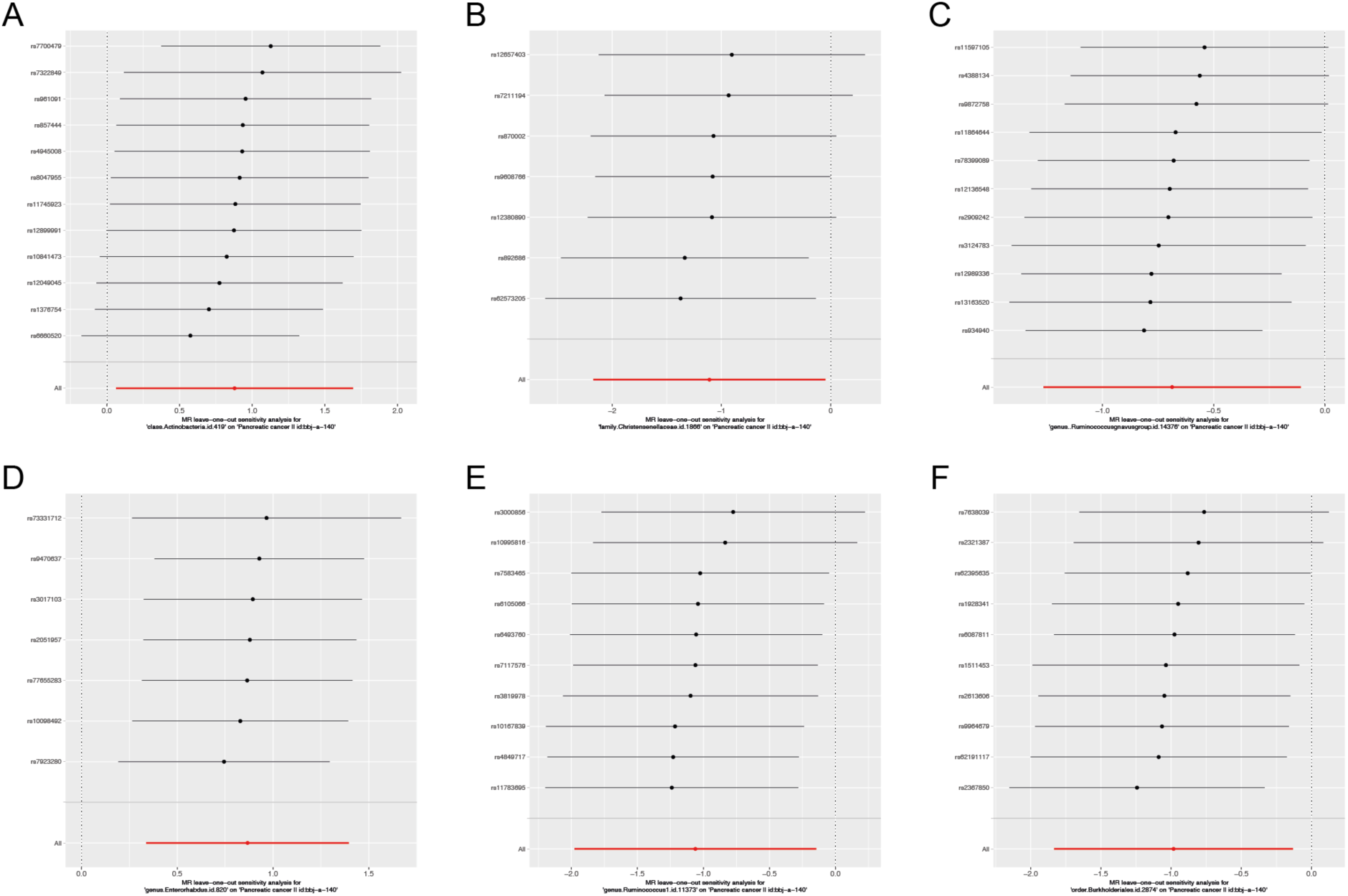
Leave-one-out sensitivity analysis for the association between genetically predicted gut microbiota and pancreatic cancer based on Japan biobank database. (A) class *Actinobacteria*; (B) family *Christensenellaceae*; (C) genus *Rumminococcusgnavus*group; (D) genus *Enterorhabdus*; (E) genus *Ruminococcus1*; (F) ordr *Burkhoderiales*

